# Sex-associated miR-4286 is related to Breslow thickness and TILs in stage II/III primary cutaneous melanoma

**DOI:** 10.64898/2026.09.04.26362289

**Authors:** Adam Z. Reynolds, David L. Corcoran, Li Luo, Klaus J. Busam, Cecilia Lezcano, Christopher Amos, Diana Argibay, Ronglai Shen, Colin B. Begg, Nancy E. Thomas, Irene Orlow, Eva Hernando, Marianne Berwick, the InterMEL Consortium

**Author notes:** Corresponding author: Adam Z. Reynolds, PhD, 2325 Camino de Salud, CRF G31, Albuquerque, NM 87131.

## Abstract

Sex differences in melanoma incidence and mortality are well-established. Previous work has shown that the female survival advantage can be largely explained by differences in clinico-pathologic features, such as age, Breslow thickness, ulceration, mitoses, and primary site. Here we test the hypothesis that sex-differentiated expression of micro-RNAs (miRNAs) may be associated with sex differences in melanoma pathology. In a sample of 715 AJCC stage II/III primary melanomas from the InterMEL project, we find two miRNAs (miR-361 and miR-4286) that are differentially expressed in males and females, and further find one of these (miR-4286) to be associated with both greater Breslow thickness and the presence of brisk tumor infiltrating lymphocytes (TILs). These results suggest that miR-4286 may be a candidate marker for sex differences in melanoma pathology. Functional experiments may confirm whether this relationship is causal or purely associative.

---

Sex disparities in melanoma incidence and mortality are robust and routinely observed. Incidence rates for men and women vary over the life course, with greater rates among women during the reproductive years until age 50, after which rates become greater among men; at all ages, however, men face worse prognosis than women and are at greater risk of dying from the disease^1^. Men tend to present with thicker, more-ulcerated tumors^2,3^, which carry more somatic mutations^4^, typically occur on the trunk (compared to the legs in women)^5^, and are more likely to recur^6,7^. One reason that men may present with more aggressive tumors is that they are less likely to engage in UV-protective behaviors, conduct self-examinations, or visit the doctor^8,9^.

Additionally, although men tend to do worse over the natural history of melanoma, male tumors are more responsive to treatment than are female tumors^10,11^, while women are at increased risk of immune-related adverse events^12^. That the female survival advantage reverses in the context of treatment is a puzzle that highlights how little is currently understood about the influence of sex on the molecular biology of melanoma.

A recent study shows that sex differences in melanoma mortality are largely mediated by the indirect effects of sex through age at diagnosis, Breslow thickness, ulceration, mitoses, and anatomical site of the primary melanoma^13^. Here we investigate whether sex-differentiated microRNAs (miRNAs) may provide one molecular mechanism for the influence of sex on these tumor features. In their mature form, miRNAs are short non-coding RNAs that regulate gene expression by binding to the 3’ UTR region of target messenger RNA (mRNA) and either repress translation or destabilize transcripts, leading to their degradation^14^. Because miRNA have short interacting (seed) regions and imperfectly bind to their targets, each miRNA can regulate dozens of potential gene targets. miRNAs have been found to exert both oncogenic and onco-suppressive effects in melanoma, associating with both clinicopathologic characteristics (e.g., Breslow thickness) and outcomes^15–23^. It has been hypothesized that miRNAs (which are disproportionately located on the X chromosome) may influence sex differences in melanoma aggressiveness or immune evasion^24,25^.

Here we use 715 early-stage primary melanomas collected as part of the InterMEL project to test the hypothesis that sex-differentiated expression of miRNAs in early-stage melanomas may contribute to sex differences in tumor pathology. The InterMEL project is a case-control study designed to investigate the natural history of untreated stage II/III primary melanoma. Eligible tumors were diagnosed between January 1998 and December 2015, and restaged according to the AJCC 8^th^ edition staging system^26^. Samples were sectioned from formalin-fixed paraffin-embedded (FFPE) tissue blocks, and nucleic acids were co-extracted using the Qiagen AllPrep DNA/RNA FFPE Kit. Laboratory protocols are presented in Orlow et al.^27^. Data are publicly available on dbGAP under accession number phs003099.v1.p1.

NanoString expression data were processed by the Hernando Lab at the New York University School of Medicine, according to methods published in their previous analysis of the same data^15^. Raw data were loaded into the R statistical programming environment^28^ and an expression threshold was determined for each sample by taking two standard deviations above the mean of the negative control probes. MiRNAs were kept for subsequent analysis if they had at least one sample above the threshold within each run date that had ten or more samples (three of the 32 run dates had fewer than 10 samples). Expression values were corrected for run date using the ComBat^29^ approach from the SVA Bioconductor^30^ package and then quantile normalized. A total of 186 miRNAs were measured and passed quality control checks. Differential expression analysis was conducted using multivariable models in the *limma* package^31^, with *p*-values adjusted using the Benjamini-Hochberg method. Breslow thickness was log-transformed. In addition to covariates shown, all models controlled for tumor purity. The voom^32^ approach was used to estimate the mean-variance trend to assign weights to observations, thereby adjusting for unequal data dispersion.

In our sample of 257 females and 458 males, nearly half (46.5%) of the males in this analysis died from melanoma within five years, while only about a third (33.9%) of females did. Males tended to be older at time of diagnosis (median: 65, IQR: 55.1–75.9) than females (median: 61, IQR: 48.5–73.5) and were more likely to have melanoma on the scalp/neck (16.0%) or trunk (33.6%), while females more often had melanomas of the lower limbs (39.8%). More males (57.6%) than females (43.4%) had at least mild/moderate solar elastosis.

Two miRNAs showed sex-differentiated expression in the InterMEL sample of stage II and III primary melanomas (Table 1; Figure 1). We found that miR-361 is down-regulated in male tumors compared to female tumors, while miR-4286 is up-regulated in male tumors (Figure 1). In models testing associations with clinico-pathologic factors, we find that miR-4286 is significantly associated with increased Breslow thickness and the presence of brisk TILs. There were also nominal associations with age, ulceration, mitoses, trunk melanomas, severe solar elastosis, and the presence of non-brisk TILs; but none of these associations were significant after adjustment for multiple tests (Table 1). Aside from its association with sex, miR-361 was not found to be associated with any of the tested factors. Neither miRNA exhibited an association with 5-year melanoma-specific mortality.

**Table 1.** Multivariable tests of sex-differentiated miRNAs in stages II/III primary melanoma. Positive (negative) log2 fold change (FC) values indicate miRNAs that are up-regulated (down-regulated) compared to the reference group. Tumor purity was also included as a covariate but is not shown here.

|  | miR-361 |  | miR-4286 |  |
| --- | --- | --- | --- | --- |
|  | Log2 FC | SD | Log2 FC | SD |
| Male<sup>a</sup> | **-0.049\*\*\*** | 0.012 | **0.072\*** | 0.010 |
| Age | -0.000 | 0.000 | -0.001 | 0.001 |
| log Breslow Thickness | -0.009 | 0.009 | **0.160\*\*\*** | 0.007 |
| Ulceration - Present | -0.007 | 0.012 | -0.034 | 0.010 |
| Mitoses - Present | 0.009 | 0.029 | -0.097 | 0.024 |
| Primary Site - Trunk<sup>b</sup> | 0.005 | 0.021 | -0.034 | 0.016 |
| Primary Site - Upper Limb<sup>b</sup> | -0.015 | 0.021 | -0.032 | 0.017 |
| Primary Site - Lower Limb<sup>b</sup> | 0.005 | 0.021 | -0.006 | 0.017 |
| Mild/Moderate Solar Elastosis | 0.018 | 0.013 | -0.014 | 0.011 |
| Severe Solar Elastosis | -0.007 | 0.021 | -0.058 | 0.017 |
| Non-brisk TILs | -0.004 | 0.020 | 0.064 | 0.016 |
| Brisk TILs | -0.029 | 0.034 | **0.255\*\*\*** | 0.026 |
| 5-year Mortality<sup>c</sup> | -0.014 | 0.013 | 0.003 | 0.011 |

Note: FDR-adjusted p-values are significant at the following levels: \* $p < 0.05$ , \*\* $p < 0.01$ , \*\*\* $p < 0.001$ .
Log2 FC = log 2-fold change. SD = standard deviation.
<sup>a</sup> Reference category is female
<sup>b</sup> Reference category is head/neck
<sup>c</sup> Reference category is survived disease free or without progression for at least five years

**Figure 1.**
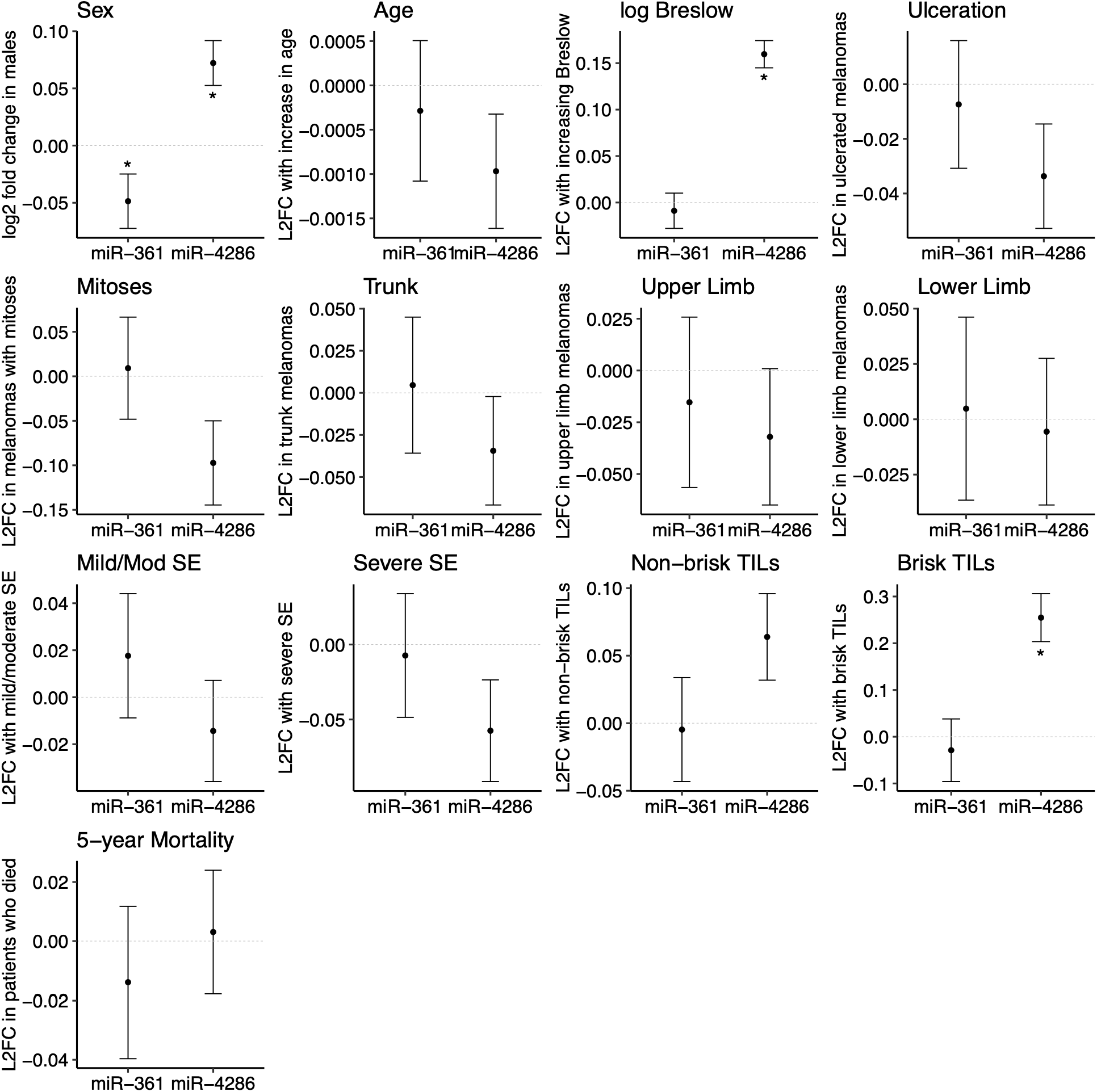
Associations between expression of miR-361 and miR-4286, sex, and clinicopathologic factors. miR-4286 is up-regulated in males and associated with both increased Breslow thickness (*p* < 0.001) and the presence of brisk TILs (*p* < 0.001). miR-361 is down-regulated in males but does not exhibit a significant association with clinicopathologic factors or mortality. Positive (or negative) Log2 fold changes (L2FC) indicate up-regulation (or down-regulation). Asterisks indicate associations that were significant after adjustment for multiple tests. Age and tumor purity were included as covariates. Reference group for primary site comparisons is head/neck. Abbreviations: SE = solar elastosis, TILs = tumor infiltrating lymphocytes.

The female survival advantage in melanoma is well-established, but the molecular mechanisms underlying it remain poorly understood. Here we find two miRNAs showing sex-differentiated expression, but neither exhibits a direct association with mortality outcomes. Our observation that miR-4286 is associated with both sex and Breslow thickness, however, suggests the intriguing possibility that it could be one molecular feature contributing to greater melanoma aggressiveness in males. Located on chromosome 8, miR-4286 has been shown to contribute to cell proliferation and inhibition of apoptosis in melanoma^33,34^, which is consistent with our finding of greater expression in thicker tumors and earlier reports showing it to be one of a number of Breslow-associated miRNAs^15^.

It is perhaps puzzling that we also find miR-4286 to be associated with brisk TIL involvement. However, this might be explained by the relationship between tumor growth and TIL recruitment. Rapidly growing tumors tend to recruit more TILs early in melanoma growth, but these TILs can vary in their immune capabilities, and thus are differently prognostic depending on tumor stage^35,36^. It is also possible that this result is a false discovery: InterMEL contains a large number of tumors with non-brisk TILs, and many fewer with brisk TILs, an imbalance which could produce a spurious association.

To our knowledge, miR-361, which is on the X chromosome, has not been functionally linked with melanoma. However, it has been identified as a tumor suppressor in other cancers, including lung, breast, ovarian, prostate, thyroid, and squamous cell carcinoma^37–42^, which is compatible with our observation of higher expression in females, who also have lower incidence and mortality from melanoma. There are several possible reasons that we did not observe miR-361 to be associated with tumor features or mortality. InterMEL consists of tumors diagnosed in stages II and III, but some molecular differences may only become relevant as the disease progresses or metastasizes. There may also be treatment effects: While the InterMEL sample consists only of early-stage patients who received no adjuvant therapy, those who later metastasized may have received treatment. Alternatively, it may be that expression of miR-361 is influenced by sex, but unrelated to disease processes in melanoma. Unfortunately, InterMEL does not include a comparison with normal tissue, and we are not aware of any publicly available miRNA datasets that are sufficiently powered to detect sex differences in healthy melanocytes.

This analysis suggests that gene dysregulation via miRNA may contribute to greater melanoma aggressiveness in males. miR-4286 is especially promising for further research into sex-specific tumor biology, to understand whether it plays a role in tumorigenesis, pathology, or poorer outcomes for males.

## Data Availability

All data are publicly available on dbGAP under accession number phs003099.v1.p1

https://dbgap.ncbi.nlm.nih.gov/beta/study/phs003099.v2.p1

## Conflict of Interest

The authors have no conflicts of interest to disclose.

## Funding

This study was supported by the following NIH grants: 1P01CA206980-01A1 to M. Berwick and N.E. Thomas; R01CA251339 to R. Shen and C.B. Begg; R01CA233524 to N.E. Thomas and C.B. Begg; R33CA160138 to N.E. Thomas and K. Conway; 5P30CA118100-15 to the University of New Mexico Comprehensive Cancer Center; P30CA016086 to the UNC Lineberger Comprehensive Cancer Center; P30CA008748 to Memorial Sloan Kettering Cancer Center; P30CA016672 to the University of Texas MD Anderson Cancer Center; P50CA225450 to I. Osman, Y. Shao, D. Polsky, and E. Hernando; P50CA221703 to J.E. Lee; R01CA121118 and R21CA245577 to S. Holmen; P30ES010126 to the University of North Carolina Chapel Hill; 1T32GM135128 to S.N. Edmiston; 1K08CA151645-01 to B.E. Gould Rothberg; R01CA112243 to N.E. Thomas, M. Berwick, and C.B. Begg.

## Data Availability

All data used in this analysis are publicly available on dbGAP under accession number phs003099.v1.p1.

## Author Contributions

Adam Z. Reynolds – Conceptualization, Formal Analysis, Writing – original draft, Writing – review & editing; David L. Corcoran – Methodology, Formal Analysis, Writing – review & editing; Li Luo – Methodology, Writing – review & editing; Klaus J. Busam – Resources; Cecilia Lezcano – Resources; Christopher Amos – Methodology, Writing – review & editing; Diana Argibay – Data curation; Ronglai Shen – Methodology, Writing – review & editing; Colin B. Begg – Conceptualization, methodology, writing – review & editing; Nancy E. Thomas – Conceptualization, supervision, project administration, funding acquisition, writing – review & editing; Irene Orlow – Conceptualization, writing – review & editing; Eva Hernando – Conceptualization, resources, supervision, project administration, funding acquisition; Marianne Berwick – Conceptualization, supervision, project administration, funding acquisition, writing – review & editing.

## Contributors

This study was conducted by the InterMEL consortium, which includes the following study centers and personnel: Coordinating Center, University of New Mexico, Albuquerque, NM, USA: Marianne Berwick (Principal Investigator (MPI, contact PI), Li Luo (Investigator), Christopher I. Amos (Advisor), Tawny W. Boyce (Data Manager), Adam Z. Reynolds (Data Analyst), Dakai Zhu (Data Analyst), Charles Wiggins (Director, New Mexico Tumor Registry); New York University, Langone Cancer Center, New York, NY: Eva Hernando (PI-Project 3), Iman Osman (co-Investigator), Douglas Hanniford (Instructor), Diana Argibay (Laboratory Technician), Una Moran (Data Technician), for the Genomics Technology Center: Adriana Heguy (Director), Sitharam Ramaswami (Associate Research Scientist); Memorial Sloan Kettering Cancer Center, New York, NY: Ronglai Shen (PI-Project 4), Colin B. Begg (Biostatistician), Arshi Arora (Biostatistician), Venkatraman Seshan (Biostatistician), Allie Reiner (Assistant Research Biostatistician), Caroline E. Kostrzewa (Assistant Research Biostatistician), Klaus J. Busam (PI-Core 2), Irene Orlow (Co-PI-Core 2), Cecilia Lezcano (Dermatopathologist), Jessica M. Kenney (Research Assistant/Sr. Laboratory Specialist), Keimya D. Sadeghi (Research Assistant and Data Engineer), Kelli O’Connell (Research Biostatistician); initial optimization assays were performed at MSK by Heta Parmar, Siok Leong, and Sergio Corrales (Research Technicians); University of North Carolina, Chapel Hill, NC: Nancy E. Thomas (MPI), Kathleen Conway (Investigator), Sharon N. Edmiston (Research Analyst), David W. Ollila (Surgical Oncologist and Investigator), Honglin Hao (Laboratory Specialist), Eloise Parrish (Laboratory Specialist), Paul B. Googe (Dermatopathologist), Stergios J. Moschos (Oncologist), David Corcoran (Bioinformatician), Lan Lin (Database Manager); Melanoma Institute Australia, Sydney, Australia: Richard A. Scolyer (Dermatopathologist and Site PI), Anne E. Cust (Epidemiologist and Site PI), James S. Wilmott (Scientist), Graham J. Mann (Cancer Geneticist and Former Site PI), Ping Shang (Scientist), Hazel Burke (Data Manager), Peter M. Ferguson (Pathologist), Valerie Jakrot (Research Manager); British Columbia Cancer Research Center, Vancouver, Canada: Tim K. Lee (Data Coordinator); Baylor College of Medicine, Houston, TX: Ivan P. Gorlov (PI-Core 3); Roswell Park Comprehensive Cancer Center, Buffalo, NY: Marc Ernstoff (Advisor), Paul N. Bogner (Dermatopathologist); The University of Texas MD Anderson Cancer Center, Houston, TX: Jeffrey E. Lee (Site PI), Melanoma Core (Technical Assistance); Dartmouth Cancer Center, Lebanon, NH: Judy R. Rees (Site PI), Shaofeng Yan (Dermatopathologist); Case Western University, Cleveland, OH: Meg R. Gerstenblith (Site PI), Cheryl Thompson (Co-Investigator); Cleveland Clinic, Cleveland, OH: Jennifer S. Ko (Dermatopathologist and Site PI), Pauline Funchain (Co-Investigator), Peter Ngo (Dermatology Fellow); Yale University Cancer Center, New Haven, CT: Marcus Bosenberg (Site PI), Bonnie E. Gould Rothberg (Former Site PI), Gauri Panse (Dermatopathologist); Columbia University Medical School, New York, NY: Yvonne M. Saenger (Site PI), Benjamin T. Fullerton (Laboratory Technician); Huntsman Cancer Institute, Salt Lake City, UT: Sheri L. Holmen (Site PI), Howard Colman (Co-Investigator), Elise K. Brunsgaard (Clinical Fellow), David Wada (Dermatopathologist); Instituto Valenciano de Oncologia, Valencia, Spain: Eduardo Nagore (Site PI), Esperanza Manrique-Silva (Dermatologist), Celia Requena (Dermatologist), Victor Traves (Pathologist), David Millan-Esteban (Post-Doctoral Fellow); Patient Advocate: Michelle Rainka.

## Notes

### Competing Interest Statement

The authors have declared no competing interest.

### Author Declarations

The IRB at University of New Mexico gave ethical approval for this work.

